# Divergent lineages of pathogenic *Leptospira* species are widespread and persisting in the environment in Puerto Rico, USA

**DOI:** 10.1101/2021.11.09.21265981

**Authors:** Nathan E. Stone, Carina M. Hall, Marielisa Ortiz, Shelby Hutton, Ella Santana-Propper, Kimberly R. Celona, Charles H.D. Williamson, Nicole Bratsch, Luis G. V. Fernandes, Joseph D. Busch, Talima Pearson, Sarai Rivera-Garcia, Fred Soltero, Renee Galloway, Jason W. Sahl, Jarlath E. Nally, David M. Wagner

## Abstract

**Background:** Leptospirosis, caused by *Leptospira* bacteria, is a common zoonosis worldwide more prevalent in the tropics. Reservoir species and risk factors have been identified but surveys for environmental sources of leptospirosis are rare. Furthermore, understanding of environmental *Leptospira* containing pathogenic genes and possibly capable of causing disease is incomplete and could result in some pathogenic strains evading detection, thereby convoluting diagnosis, prevention, and epidemiology.

**Methodology/Principal Findings:** We collected environmental samples from 22 sites in Puerto Rico during three sampling periods over 14-months (Dec 2018-Feb 2020); 10 water and 10 soil samples were collected at each site. Samples were screened for pathogenic *Leptospira* DNA using the *lipL32* PCR assay and positive samples were sequenced to assess genetic diversity. One urban site in San Juan was sampled three times over 14 months to assess persistence in soil; live *leptospires* were obtained during the last sampling period. Isolates were whole genome sequenced and LipL32 expression was assessed *in vitro*.

We detected pathogenic *Leptospira* DNA at 15/22 sites; both soil and water were positive at 5/15 sites. We recovered *lipL32* sequences from 83/86 positive samples (15/15 positive sites) and *secY* sequences from 32/86 (10/15 sites); multiple genotypes were identified at 12 sites. These sequences revealed significant diversity across samples, including four novel *lipL32* phylogenetic clades. Most samples from the serially sampled site were *lipL32* positive at each time point. We sequenced the genomes of six saprophytic and two pathogenic *Leptospira* isolates; the latter represent a novel pathogenic *Leptospira* species likely belonging to a new serogroup.

**Conclusions/Significance:** Diverse and novel pathogenic *Leptospira* are widespread in the environment in Puerto Rico. The disease potential of the novel lineages is unknown but several persisted for >1 year in soil, which could contaminate water. This work increases understanding of environmental *Leptospira* and should improve leptospirosis surveillance and diagnostics.

**Author Summary:** Leptospirosis is a common zoonotic disease worldwide, but more prevalent in the tropics. Cases are more common following severe weather events, possibly due to flooding, which may more readily distribute soil and/or water contaminated with *Leptospira* spp., the disease agents. Human cases increased following the 2017 hurricanes that ravaged Puerto Rico (Maria and Irma), prompting environmental sampling of soil and water to assess the presence, abundance, and persistence of pathogenic *leptospires* in these environments. The goal was to better understand these potential reservoirs of human and animal disease. Divergent and novel groups of pathogenic *Leptospira* were abundant and widespread in soil and water in Puerto Rico and sometimes persisted in these environments for >1 year. However, most groups we identified have not previously been described from humans and/or other animals, so the disease potential of these novel organisms is unknown. The results of this study reveal a tremendous amount of previously uncharacterized *Leptospira* diversity in soil and water in Puerto Rico, which could contribute to cryptic disease. The description and characterization of these novel types improves our understanding of the genus *Leptospira*, and will aid in the developent of improved diagnostics and preventative tools to advance public health outcomes.

## Introduction

Leptospirosis is a one-health zoonotic disease of worldwide importance (1) and the most widespread zoonosis globally, affecting a wide variety of mammalian species (2, 3). It is caused by pathogenic bacteria of the genus *Leptospira*. There are three broad groups of leptospires within the genus that are categorized based upon 1) phylogenetic similarities, 2) *in vitro* phenotypes, and 3) actual or presumed pathogenicity (i.e., the presence of the lipL32 pathogenicity gene) (1, 4). To date, across four groups, there are 17 described species in the pathogenic group (group P1), 21 species in the intermediate group (P2), and 26 in the saprophytic groups (S1 and S2) (1).

Leptospires are obligate aerobic spirochetes that are highly motile and slender (0.15 × 10–20μm) (5). Their genomes are comprised of two circular chromosomes and are larger than other spirochetes and more diverse than those of many other bacterial genera (6) due to horizontal gene transfer and gene duplication (7). This potentially facilitates an increased ability to survive in a variety of hosts (humans, domestic and wild animals), environmental conditions (soil and water), and climates (tropical, temperate, etc.) (6, 8).

Leptospirosis is maintained in animal reservoir hosts, such as rodents, cattle, and domestic dogs (9, 10), with humans serving as incidental hosts. Leptospires colonize the proximal renal tubules of infected animals and these reservoir hosts then excrete leptospires in their urine, thereby contaminating the environment wherein leptospires can survive under moist conditions (11) and infect other susceptible hosts (1, 12). Transmission of leptospirosis typically occurs via direct contact with infected urine or indirectly through exposure to contaminated soil and water (12). Leptospirosis infections in animals may become chronic and can last for months or years (13). During chronic infection the shedding of leptospires in urine is intermittent (14), which convolutes the identification of infected animals and disease mitigation efforts.

Leptospirosis is a major global public health concern that disproprotionately affects resource-poor populations (15), especially developing countries with poor sanitation and dense urban centers where rats, a common reservoir host, are abundant (12). Thus, people living in urban centers and those directly exposed to infected livestock animals seem to be at highest risk for leptospirosis, probably due to increased exposure rates (16). Recent estimates of global human disease cases are 1.03 million annually, including 58,900 deaths (15); 73% of cases occur in the tropics (17).

In the United States (US), leptospirosis is considered endemic with most human cases arising in Puerto Rico and Hawaii (17). Leptospirosis was first reported in Puerto Rico in 1942 (18) and it is common in other parts of the Caribbean: 12,475 human cases were reported by nine Caribbean countries between 1980 and 2005 (19). Despite this, it was removed from the US reportable disease list in 1995 but later reinstated in 2014. During this time (1995-2014) and despite the lack of requirement to report this disease, 759 cases of leptospirosis were reported in Puerto Rico; 570 of these were confirmed, including 92 deaths. A retrospective analysis of these cases by Santiago-Ramos *et al*. (18) identified a trend between annual case rates and annual precipitation, although case rates were homogenous across Puerto Rico (*i*.*e*., geographical hot spots were not identified). Most cases occurred in males and in densely populated urban areas (18).

Heavy rains and floods have been identified as major risk factors for human leptospirosis, particularly in tropical regions and on islands (20). Hurricanes are associated with leptospirosis outbreaks (17, 21–23), and the severity of these major storms can be amplified due to conditions associated with climate change (e.g., increased ambient temperatures and humidity). Hurricane Hortense in 1996 (24) and the 2017 hurricanes Irma and Maria caused major damage to infrastructure across Puerto Rico, which limited access to potable water, as well as electricity, communications, and transportation (25). In just the three months following hurricanes Maria and Irma (26), the US Centers for Disease Control and Prevention (CDC) identified 85 leptospirosis cases, including 14 deaths, which was greater than the reported annual cases for any of the three previous years (2014-2016; 45-73 annual cases) (CDC personal communication).

Although some current thinking suggest pathogenic leptospires are unable to multiply in the environment (27, 28), leptospirosis has nonetheless been considered an environmentally-borne infection (29) because indirect contamination via the environment is the most frequent source of human infection (28). However, in recent years the use of the term “environmental reservoirs” for leptospirosis has increased, particularly when referring to moist soil in endemic regions (28), and the discovery of novel, pathogenic leptospires persisting in these environments (this study and (1)) supports the adoption of this term. Although moist soil conditions seem to be associated with environmental persistence of leptospires (30–32) little is known about the duration of survival in the environment (28, 33). Environmental surveys are thus of critical importance toward our understanding of these environmental reservoirs and their role in the persistence and proliferation of pathogenic leptospires, as well as their potential role in disease transmission. To this end, we conducted environmental surveys in Puerto Rico after the two 2017 hurricanes to: 1) characterize the prevalence and geographic distribution of pathogenic *Leptospira* spp. in soil and water, 2) understand the diversity of pathogenic strains that occupy these environments, 3) investigate persistence of pathogenic *Leptospira* spp. in moist soil, and 4) isolate and characterize pathogenic *Leptospira* spp. persisting in soil in Puerto Rico.

## Methods

### Sample collection

The focus of our environmental sampling efforts, conducted over three time periods (Dec 3-14, 2018; Feb 9-23, 2019; and Feb 13-14, 2020), were 12 municipalities in Puerto Rico (**Figure 1**) in which there is a potentially higher leptospirosis risk to humans based on previous human and canine leptospirosis cases and environmental factors, such as areas prone to flooding and poor sanitation (18). We collected 440 samples (10 water and 10 soil per site) from 22 field sites within these municipalities (**Table S1**); we targeted urban rivers and canals as well as runoff sites and/or flood zones near livestock farms where urine contamination from livestock was likely. At each site we established five short (<3m) linear transects and collected two soil samples (~50g each) and two water samples (>150mL each) per transect (sampling strategy illustrated in **Figure S1**). For each transect, the first soil sample was collected from the edge of the water and the second soil sample was 1m back and directly perpendicular to the edge of the water. Likewise, the first water sample was collected at the edge of the water and the second water sample was collected 1m beyond and perpendicular to the edge of the water (**Figure S1**). Soil samples were collected ≤10cm below the surface and placed into 50mL capacity conical tubes and, when possible, water samples were collected from stagnate or less turbulent areas of the subsurface water (10-30cm deep) into a Whirl-Pak. For site 22, collection in transects was not possible due to access constraints at this urban canal. At that site we collected 10 water and 10 soil samples within ≤1m of the edge of the water. For soil samples at all sites we recorded approximate elevation of the samples relative to the water, in meters. Soil elevation was categorized into four groupings: 0-0.33m, >0.33-0.66m, >0.66-1m, and >1m (**Table S2**). Finally, we revisited one pathogenic Leptospira spp. positive site in San Juan (site 16) that was identified during collection period 1 (Dec 3-14) twice more, once in February 2019 and again in February 2020 (**Figure 1**). In February 2019 we collected 10 additional soil samples at this site in a linear transect along the edge of the water in the same location where the previous positive soil samples were collected. In February 2020, for the purpose of culturing attempts to obtain live leptospires, we collected 20 soil samples at this site clustered at the edge of the water (two clusters of 10 samples) where we detected a high abundance of pathogenic *Leptospira* DNA from the previous two collection periods.

**Figure 1:**
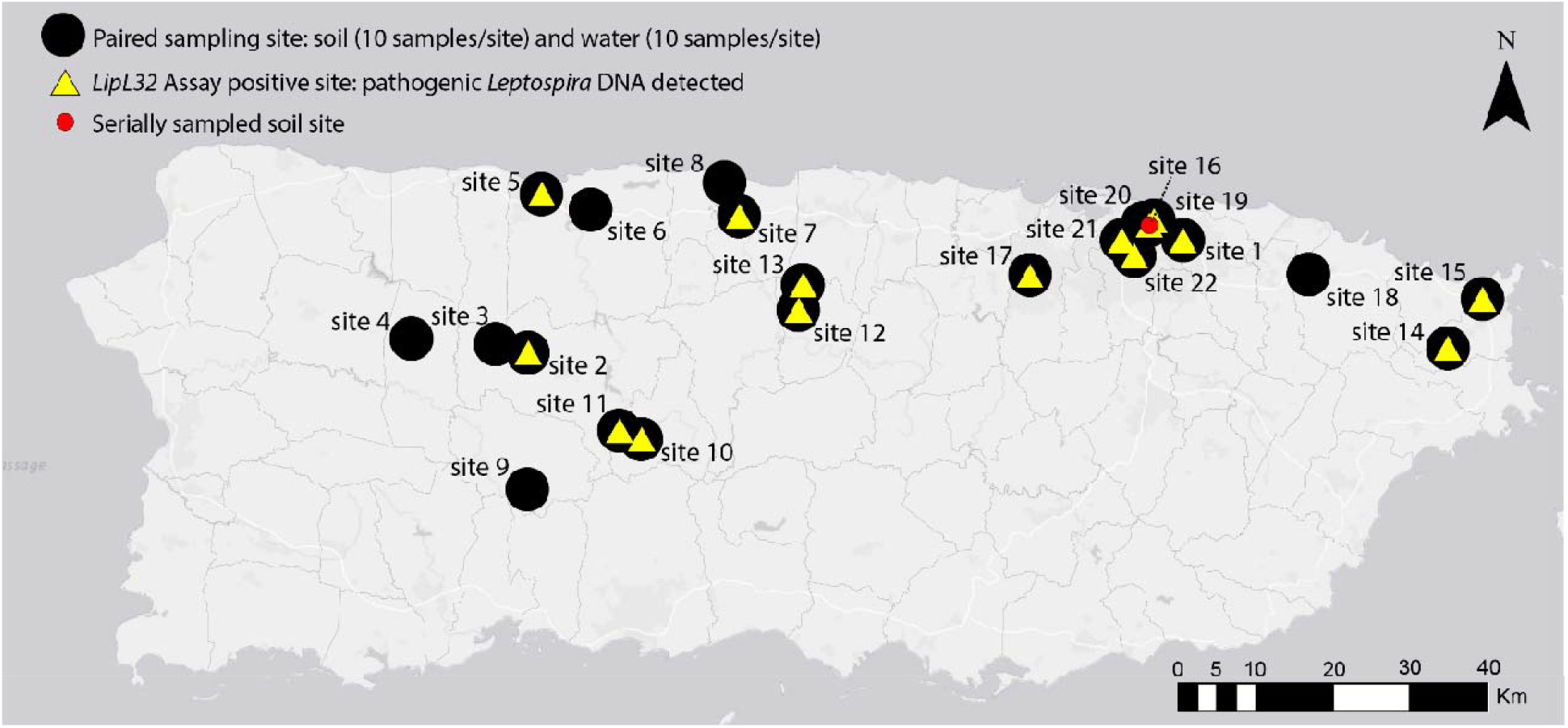
Map of Puerto Rico indicating sites where environmental samples were collected for the detection of pathogenic *Leptospira* spp. DNA from 2018-2020. Black dots represent paired water and soil sites (n=22); 20 samples were collected from each site (10 soil and 10 water). Yellow triangles indicate sites where pathogenic *Leptospira* DNA was detected (15/22 sites). One site in San Juan (site 16, red circle) was serially sampled three times over a 14-month period to assess persistence of pathogenic *Leptospira* in soil.

### Soil and water processing

Water samples (150mL) were filtered through a 250mL capacity 0.22µm nitrocellulose filter as previously describe (34); afterward the filter was cut in half using sterile scissors and half of the filter was placed in a 5mL DNeasy® PowerWater DNA bead tube (Qiagen, Valencia, CA, USA) and refrigerated at 4°C until shipment to the laboratory. Prior to filtering, 1mL of each homogenous water sample was placed in a 2mL capacity screw-cap tube for downstream pH testing. We collected pH values directly from the water sample using a 6mm glass pH probe (Cole-Parmer, Vernon Hills, IL, USA, Part # EW-55510) and from soil using methods outlined by Hall *et al*., 2019 (34) (**Table S2**).

### DNA extractions

DNA extractions were conducted in a class II A2 biosafety cabinet (BSC) as required by our USDA-APHIS-PPQ soil transport permit. DNA was isolated from water filters using DNeasy® PowerWater DNA extraction kits (Qiagen, Valencia, CA, USA) following the recommendations of the manufacturer, and from ~0.5g of soil using Qiagen PowerSoil DNA extraction kits (Qiagen, Valencia, CA, USA) following previously published conditions (35) with one modification: after the addition of solution C3, the sample was incubated at 4°C overnight. DNA extractions were assessed for overall quality and bacterial abundance using a universal 16S real-time PCR assay as previously described (35); this quality control step is necessary prior to conducting a pathogen detection PCR assay because it provides confidence that a negative result truly reflects the absence of pathogen DNA and not a technical failure associated with poor quality or low yield DNA extracts.

### *Leptospira* qPCR detection and direct sequencing

All DNA extracts were screened in triplicate 10µL PCRs to detect the presence of DNA from pathogenic *Leptospira* spp. using the *lipL32* TaqMan® real-time PCR assay (4) containing the following: 1x TaqMan® Environmental PCR master mix (Applied Biosystems, Foster City, CA, USA), 0.9µM of each primer, 0.45µM of the MGB probe, and 1µl undiluted DNA template. Quantitative PCRs were run interchangeably on Applied Biosystems QuantStudio® (QS) QS7 and QS12 Real-Time PCR Systems with QS 12K Flex or QS Real-Time PCR software, as appropriate, under the following conditions: 50°C for 2 minutes, 95°C for 10 minutes, and 45 cycles of 95°C for 15 seconds and 58°C for 1 minute. Positive (*L. interrogans* strain Fiocruz L1-130) and non template controls were included on all runs. *LipL32* amplicons from all positive samples were subjected to direct Sanger sequencing to confirm that the amplified product was *Leptospira*, using the same forward and reverse primers for the PCR (all replicates from a single sample were sequenced independently). Treatment and sequencing conditions are described below.

### secY 203bp amplification

All samples that yielded a positive PCR result with the *lipL32* assay (defined as amplification of at least 1 of 3 replicates with a C_t_ <45) were also subjected to PCR amplification of the secY gene; we amplified a 203bp fragment using the “SecYIVF” and “SecYIVR” primers designed to amplify pathogenic *Leptospira* spp. as described in Ahmed *et al*., 2009 (36). PCRs were carried out in 10µL volumes containing the following reagents (given in final concentrations): 1µL of 1/25 diluted DNA template (none of the PowerWater® extractions were diluted), 1x SYBR® Green Universal master mix (Applied Biosystems, Foster City, CA, USA), 1.2M betaine, 0.2U Platinum® Taq DNA polymerase (Invitrogen, Carlsbad, CA, USA) to improve efficiency, and 0.4µM of each primer. The assay was run on an Applied Biosystems 7500 Fast Real-Time PCR System with SDS 7500 software v2.0.6 under the following conditions: 50°C for 2 minutes, 95°C for 10 minutes, and 40 cycles of 95°C for 15 seconds and 57°C for 1 minute. Positive and non template controls were included on all runs. Due to the low abundance of *Leptospira* DNA in many of these samples, triplicate reactions were employed to increase the number of *secY* sequences generated; frequently only a single reaction (out of three) would yield an amplicon.

### *secY* 549bp amplification

We also attempted to sequence a 549bp region of the *secY* gene for all *lipL32* positive samples to gain more phylogenetic resolution for certain samples and increase the number of *secY* sequences generated for this dataset. We used primer pair F-ATGCCGATCATTTTTGCTTC and R-CCGTCCCTTAATTTTAGACTTCTTC, which were designed to amplify pathogenic *Leptospira* spp. as previously described (37). Duplicate PCRs were carried out in 10µL volumes containing the following reagents (given in final concentrations): 1µL of 1/25 diluted DNA template (undiluted for all PowerWater extractions), 1x PCR buffer, 2.5mM MgCl2, 0.2mM dNTPs, 0.8U Platinum® *Taq* DNA polymerase, and 0.4µM of each primer. PCRs were thermocycled according to following conditions: 95°C for 10 minutes to release the polymerase antibody, followed by 38 cycles of 94°C for 60 seconds, 55°C for 30 seconds, and 72°C for 30 seconds, and a final extension step of 72°C for 10 minutes to ensure completion of the fragments. Positive and non template controls were included on all runs.

### Sanger sequencing

For all three amplicons (*lipl32*-242bp, *secY*-203bp, and *secY-*549bp), PCR products were visualized on a 2% agarose gel to ensure that the products were of the expected size and treated with 1µL of ExoSAP-IT (Affymetrix, Santa Clara, CA, USA) added to 7µL of PCR product under the following conditions: 37°C for 15 minutes followed by 80°C for 15 minutes. Treated products were then diluted (based on amplicon intensity) and sequenced in both directions using the same forward and reverse primers from the PCR in a BigDye® Terminator v3.1 Ready Reaction Mix (Applied Biosystems, Foster City, CA, USA). We used 10µL volumes for sequencing reactions containing the following reagents (given in final concentrations): 5x Sequencing Buffer, 1µL BigDye® Terminator v3.1 Ready Reaction Mix, 1µM primer, and 5µL diluted PCR product. The following thermocycling conditions were used: 96°C for 20 seconds, followed by 30 cycles of 96°C for 10 seconds, 50°C for 5 seconds, and 60°C for 4 minutes. Sanger sequences were assembled and primers were manually removed using SeqMan Pro (DNASTAR Lasergene, Madison, WI, USA), resulting in sequence lengths of 202bp (*lipL32*-242bp), 162bp (*secY-*203bp), and 504bp (*secY*-549bp). FASTA database files for each gene *(lipL32* and *secY*) were generated in BioEdit (38); sequences from both secY amplicons (162 and 504bp) were combined into a single FASTA file.

### *lipL32* high fidelity amplicon sequencing

Sanger sequences from a subset of samples displayed multiple genotypes from replicate amplicons and/or heterogeneous nucleotide calls in a single sequence, suggesting multiple *Leptospira* spp. genotypes (*i*.*e*., mixtures). Also, within some *lipL32* positive samples the concentration of *Leptospira* spp. DNA was too low to sequence using traditional Sanger sequencing methods. Thus, to recover sequences from these low-level positives and identify and characterize individual genotypes within these mixtures, we employed a high fidelity amplicon sequencing approach (AmpSeq) that enables differentiation of alleles without the need for traditional cloning and sequencing of the PCR amplicon while providing confidence in the detection and sequencing accuracy of low level positives (39). We amplified the same 242bp fragment of the *lipL32* gene described above in triplicate reactions using modified primers that incorporated universal tails (39). The PCR setup and conditions for this modified *lipl32* AmpSeq assay were the same as described above. Replicates were pooled and sequence libraries were prepared for all positive samples as previously described (39), except a 1x Agencourt AMPure XP (Beckman Coulter, Indianapolis, IN, USA) bead cleanup was used. Uniquely indexed sample libraries were pooled together in equimolar amounts and sequenced on an Illumina MiSeq instrument using a 500 cycle (2 × 250) MiSeq Reagent Kit v2 Nano with PhiX control (Illumina, San Diego, CA, USA, part# MS-103-1003). We sequenced all lipL32 positive soil and water samples (n=86) in parallel on a single run.

### Illumina sequence processing and *ipL32* phylogenetic analysis

Paired-end Illumina reads were processed within QIIME 2 (v2019.7 and 2020.6) (40); primers were trimmed with cutadapt (41). The DADA2 (42) QIIME 2 plugin was used for quality trimming (--p-trunc-len-f 115 --p-trunc-len-r 110), denoising, dereplication of sequences, and chimera removal. *LipL32* sequences were identified with BLASTN (43) searches of representative sequences against the NCBI nr/nt database (NCBI Resource Coordinators 2016) and BLAST databases consisting of *lipL32* Sanger sequences generated from *Leptospira* positive samples as part of this study. *LipL32* Illumina and Sanger sequences were combined with 538 publically available *lipL32* sequences that were extracted from whole genomes (GenBank accession numbers in **Table S3**) using BLASTN v2.9.0+ (44) (*lipL32* sequence from *L. interrogans* strain Fiocruz L1-130 was used as a reference) and aligned with MUSCLE (v3.8.31) (45). Phylogenetic analysis was conducted using MEGA version 7 (46) and a maximum likelihood phylogeny was inferred using the Tamura-Nei model; bootstrap values were calculated using 1,000 replicates.

### *secY* phylogenetic analysis

Near full-length *secY* gene sequences (~1,300bp) were extracted from publicly available genome assemblies (**Table S3**) with BLASTN (v2.2.29) (47). These nucleotide sequences were combined with Sanger sequences generated from environmental samples in this study (162bp and 504bp amplicons) and aligned with MUSCLE (v3.8.31) (45). A maximum likelihood phylogeny was inferred with IQ-TREE (v1.6.12) (48) using the best-fit model (TIM+F+I+G4) identified by ModelFinder (49) and the UFBoot2 ultrafast bootstrapping (50) and SH-aLRT (51) options.

### Statistical analyses

We tested for associations between environmental and sampling variables and the detection of pathogenic *Leptospira spp*. in soil and water (**Table S2**). Sampling variables were not available for 13 soil samples and site 22 was removed entirely from this analysis because the sampling strategy for this site was different from the other sites (see above). Sample sizes used for each test are listed in **Table S4**. For categorical variables (sample type, location, and elevation), we used chi-square tests of independence, whereas for pH (a continuous variable) we used Wilcoxon Rank Sum (W) test because these data were not normally distributed. Categories for each environmental and sampling variable are described above under “Sample collection” and “Soil and water processing” and listed in **Table S2;** p-values of <0.05 were considered significant. All analyses were conducted in R studio v3.6.1 with tidyverse and ggplot2 packages (52).

### Soil culturing

Ten grams of soil samples were resuspended in 20mL of sterile water in 50mL conical tubes, vigorously vortexed, and allowed to settle for 15 minutes for sedimentation of solid matter. Supernatants were filtered through a 0.45µm filter as a selection step to remove larger bacteria, and 2mL of filtrate was used to inoculate either a 2x HAN (53) or 2x EMJH media (Difco, BD, Franklin Lakes, NJ, USA), both containing STAFF (54). EMJH cultures were grown at 29°C and HAN cultures were prepared in duplicate for growth at both 29 and 37°C. Each culture was assessed daily by dark-field microscopy and, upon reaching high densities of growth (>10^8^ spirochetes/mL), an aliquot was removed for flourescent antibody testing prior to storage in liquid nitrogen for downstream analysis.

### Fluorescent Antibody Test (FAT)

Ten microliters of cultured samples were placed on a glass slide with a 7mm well as previously described (55). Briefly, slides were air dried overnight and fixed in acetone for 15Lminutes and then placed in a humid chamber; 50μL of rabbit anti-LipL32 sera (1:250) was added to each spot and then incubated for 1 hour at 37°C. Next, slides were washed for 10⍰minutes in PBS with gentle rocking and incubated with secondary Alexa Fluor 488 F(ab’)2 goat anti-rabbit IgG (Invitrogen, Waltham, MA, USA) for 1 hour at 37°C in the dark. After extensive washing, slides were dried and counterstained for 15⍰seconds with Flazo Orange (1:50, National Veterinary Services Laboratory). Slides were then rinsed with PBS and mounted using ProLong™ Diamond Antifade Mountant with DAPI (Thermo Fisher, Waltham, MA, USA). Microscopic examination was done using a Nikon Eclipse E800 microscope and B2-A filter (excitation, 450–490⍰nm; emission, 520⍰nm) at 400× magnification (56).

### ELISA

Cultured spirochetes from soil were recovered from liquid media (4,000 x g for 15 minutes), washed twice with PBS, and resuspended in 500µL of PBS. Cells were counted by dark field microscopy, suspensions were brought to a concentration of 5×10^7^ spirochetes/mL, and 100µL (~5×10^6^) was used to coat individual wells on ELISA plates in triplicate (Thermo Fisher, Waltham, MA, USA) for 16 hours at room temperature. Plates were then washed three times with PBS and blocked with PBS-containing 1% bovine serum albumin (BSA) for 1 hour at 29°C. For detection of pathogenic leptospires within the sample population, wells were incubated for 1 hour with polyclonal rabbit anti-LipL32 (1:1,000 in blocking buffer) followed by secondary horseradish peroxidase-conjugated goat anti-rabbit IgG (1:3,000). Wells were washed six times and reactivity was revealed by the addition of 100µL of TMB substrate (SeraCare, Milford, MA, USA) for 15 minutes. Reactions were stopped with 50µL of TMB STOP™ Solution and optical densities were taken at 620nm in a 96-well plate reader. Sample analysis was performed in triplicate. Positive and negative controls were comprised of wells coated with 100µL (containing 5×10^6^, 2.5×10^6^, 1.25×10^6^, or 6.25×10^5^) of L. interrogans serovar Copenhageni (LipL32 positive) and 100µL (5×10^6^) *L. biflexa* serovar Patoc (LipL32 negative), respectively.

### Selection and culture of pathogenic leptospires

ELISA and FAT-positive mixed cultures were enumerated by dark field microscopy and leptospires were diluted in HAN media to 10^4^ cells/mL; 100µL was spread onto HAN agar plates supplemented with 0.4% rabbit serum (56) and incubated at 37°C until colonies were visible. Individual colonies were then harvested from the plates, vigorously homogenized in 100µL of HAN for microscopic visualization and, after confirmation of motile leptospires, 10µL of each suspension was used for coating FAT slides for LipL32 detection, as described above. The remaining sample was maintained at 29°C and, in the event of a positive FAT with anti-LipL32, used to inoculate fresh HAN media. Cultures were grown at 37°C, harvested at mid-log phase, and were further confirmed as pathogens by PCR with the *lipL32* and *secY*-549bp primer sets described above. *SecY* amplicons were Sanger sequenced (as above) to identify and differentiate individual colonies.

### WGS sequencing of isolated leptospires

Genomic DNA was extracted from isolated colonies using DNeasy kits (Qiagen, Valencia, CA, USA) according to the recommendations of the manufacturer, except the buffer AL incubation step occurred at 80°C for 1 hour. The gDNA was assessed for quality and quantity on a 0.7% agarose gel using λ DNA-HindIII Digest (New England Biolabs, Ipswich, MA, USA). WGS library construction was performed using the KAPA Hyper Prep Kits for Illumina NGS platforms per the manufacturer’s protocol with double-sided size-selection performed after sonication (KAPA Biosystems, Woburn, MA, USA, part# KK8504). The adapters and 8bp index oligos purchased from IDT® (Integrated DNA Technologies, San Diego, CA, USA), based on Kozarewa and Turner, 2011 (57), were used in place of those supplied in the KAPA preparation kit. The final libraries were quantified on an Applied Biosystems™ QuantStudio™ 7 Flex Real-Time PCR System using the KAPA SYBR® FAST ROX Low qPCR Master Mix for Illumina platforms (part# KK4873). The libraries were then pooled together at equimolar concentrations and quality was assessed with a Bioanalyzer High Sensitivity DNA kit (Agilent Technologies, Santa Clara, CA, USA, part# 5067-1504). Final quantification by qPCR preceded sequencing of the final library. The samples were sequenced on an Illumina MiSeq using the 600-cycle v3 kit (part# MS-102-3003) with the standard Illumina procedure. The appropriate sequencing primers were added to the MiSeq kit as previously described (57).

### WGS phylogenetic analysis

#### External genomes

Publically available *Leptospira* genomes were downloaded from the GenBank assembly database (58) with the ncbi-genome-download tool (https://github.com/kblin/ncbi-genome-download) on September 9th, 2021. Genomes were renamed to include the assembly accession and taxonomic information. The dataset resulted in a set of 802 reference genomes that included pathogenic, intermediate, and saprophytic leptospires (**Table S3**).

### Genome assembly

Raw reads were trimmed with bbduk.sh (v38.92) (https://sourceforge.net/projects/bbmap/) and assembled with SPAdes (v3.13.0) (59). Reads were mapped back against contigs with minimap2 (v2.22) (60) and the depth of coverage was calculated with Samtools (v1.13) (61). Two hundred nucleotides from each contig were aligned against the GenBank nt database with BLASTN (v.2.11.0) (44) and the taxonomy of the top hit was recorded. Contigs with an anomalously low depth of coverage or aligned against contaminants were manually removed.

#### Leptospira dendrogram

Pairwise MASH distances (62) were calculated on all *Leptospira* genomes and a dendrogram was generated with mashPy (https://gist.github.com/jasonsahl/24c7cb0fb78b4769521752193a43b219), a tool that incorporates SciPy (63) and Skbio (http://scikit-bio.org).

### Average nucleotide identity

Average nucleotide identity (ANI) between the novel *Leptospira* spp. described herein and *L. yasudae* strain 201601115 was calculated using the ANIb stat in PYANI (v0.2.11) as described previously (64)

### SNP and indel discovery

Genome assemblies were annotated with Prokka (v1.14.6) (65). Reads were aligned against each reference and SNPs and indels were called with Snippy (v4.6.0) (https://github.com/tseemann/snippy) using default parameters.

### Core genome SNP phylogenies

For pathogenic genomes, assemblies were aligned against *Leptospira kmetyi* strain LS-001/16 (GCA_003722295.1) with NUCmer (v3.1) (66) and SNPs were called with NASP (v1.2.0) (67); SNPs that fell within duplicated regions, based on a reference self-alignment with NUCmer, were filtered from downstream analyses. A maximum likelihood phylogeny was inferred on the concatenated SNP alignment with IQ-TREE (v2.0.3) (68) in conjunction with ModelFinder (49) and rooted with *Leptospira interrogans* FDAARGOS 203 (GCA_002073495.2). The reference genome, *Leptospira montravelensis* strain 201800278 (GCA_004770045.1), was used for the phylogeny including saprophytic genomes, which was rooted by *Leptospira noumeaensis* strain 201800287 (GCA_004770765.1).

### Hamster infection with isolated colonies

To assess virulence of the newly identified pathogenic isolate described herein, Golden Syrian hamsters were inoculated by intraperitoneal (IP) injection with 10^8^ leptospires in 1mL as previously described (69). Negative control hamsters received 1mL of media alone. Animals were assessed for acute disease through day 11 post infection and euthanized on day 21 to provide ample time for renal carriage to develop. All animal experimental procedures were performed in accordance with relevant guidelines and regulations, and as approved by USDA institutional guidelines.

### Microscopic agglutination test (MAT)

The MAT was performed according to OIE guidelines (70) at two-fold dilutions from an initial dilution of 1:50 to 1:6400 using 18 serovars of *Leptospira* spp. listed in **Table S5** and the newly identified pathogenic isolates described herein.

### Culture

At three weeks post-infection, kidneys were harvested using aseptic techniques for culture of leptospires in T80/40/LH semi-solid media incubated at 29°C (71) and HAN semi-solid media incubated at both 29 and 37°C (53).

### Data sharing

All WGS reads and assemblies generated during this study have been deposited in NCBI BioProject database under accession # PRJNA766613. The associated BioSample accession numbers for these isolates are sequentially assigned beginning with # SAMN21833208 and ending with # SAMN21833215. Likewise, SRA accession numbers for these isolates are sequentially assigned beginning with # SRR16134453 and ending with # SRR16134460. Partial gene sequences for *lipL32* and *secY* have been deposited in NCBI (accession numbers sequentially assigned beginning with # OK345072 and ending with # OK345268) and are also provided in Supplemental File 1 (**Table S6 and Table S7**).

## Results

### Pathogenic *Leptospira* detection

Pathogenic *Leptospira* spp. were common and distributed widely throughout Puerto Rico. Using *lipL32* PCR, we detected pathogenic *Leptospira* spp. DNA at 15 of 22 paired soil and water exploratory sites (68.2%) (**Figure 1**); 8/15 sites yielded positive soil samples only, 2/15 positive water samples only, and 5/15 both positive soil and water samples (**Figure S2**). From these exploratory sites, 63 samples (out of 440) were positive, including 15 water and 48 soil samples. At the serially sampled soil site (site 16), an additional 23 (out of 30 total) soil samples collected during the second and third visits to this site also were positive. In total, 86 positive samples from Puerto Rico were detected. At five paired soil and water sites (sites 7, 12, 17, 21, and 22) we detected pathogenic *Leptospira* spp. in both sample types (**Table 1** and **Figure S2**). At four of these five sites (12, 17, 21, and 22) a higher proportion of soil samples were positive than water samples and the phylogenetic clades represented in the water samples were a subset of those identified in the corresponding soil samples (**Table 1**). This observation was not possible at site 7 because only a single soil and a single water sample were *lipL32* positive and *lipL32* sequence was not recovered from the positive soil sample (**Table S1**).

**Table 1:** Table of all positive sites in Puerto Rico illustrating the relationship between *lipL32* genotypes found in soil and water. In total, 15 sites were positive for pathogenic *Leptospira* DNA; eight sites wherein only soil was positive, two sites wherein only water was positive, and five sites wherein both soil and water were positive (highlighted in gray). At four of the latter sites, the clades present in water were a subset of the clades identified in soil (bolded text), suggesting that soil reservoirs may lead to the contamination of water.

| Site ID | Site Type | Proportion Positive | Clades |
| --- | --- | --- | --- |
| 1 | Water | 0.1 | 4b |
| 2 | Soil | 0.2 | 4a |
| 5 | Water | 0.7 | 1, 6 |
| 7 | Water | 0.1 | 6 |
| 7 | Soil | 0.1 | unknown |
| 10 | Soil | 0.1 | 4a |
| 11 | Soil | 0.2 | 4a |
| 12 | Water | 0.1 | **4b** |
| 12 | Soil | 0.6 | 3, 4a, **4b**, 6, 9 |
| 13 | Soil | 0.1 | 6 |
| 14 | Soil | 0.4 | 3, 4a, 4b, 9 |
| 15 | Soil | 0.3 | 4b |
| 16 | Soil | 0.7 | 4b, 6 |
| 17 | Water | 0.1 | **6** |
| 17 | Soil | 0.4 | 3, 4b, **6** |
| 19 | Soil | 0.3 | 4b, 6 |
| 21 | Water | 0.1 | **6** |
| 21 | Soil | 0.9 | 4a, 4b, **6**, 9 |
| 22 | Water | 0.3 | **4b**, **6** |
| 22 | Soil | 0.5 | **4b**, **6**, 9 |

### Gene sequencing *lipL32*

Sequencing of the *lipL32* amplicons revealed extensive and previously undescribed diversity in pathogenic *Leptospira* spp. at this locus. Using AmpSeq and Sanger sequencing methods, we generated 147 *lipL32* sequences from 83 of the 86 positive samples representing all 15 positive sites (**Table S1**) to: 1) confirm that the generated amplicons were from *Leptospira* DNA, and 2) assess genetic diversity of pathogenic *Leptospira* spp. within and across sites and samples; information on generated nucleotide sequences is provided in **Table S6**. Thirty-four alleles were identified in this dataset from Puerto Rico of which 32 were novel *(i*.*e*., were not a 100% nucleotide match to any available sequences in GenBank). We assigned either a high or moderate level of confidence to each newly identified allele. If a sequence was observed in more than one sample or was generated via Sanger sequencing methods, this allele was assigned a high level of confidence; 20 of 34 alleles met this criterion. The remaining 14 alleles were only observed in a single sample, only in the Illumina dataset, and/or with a low depth of coverage (3-42 paired end reads) and thus were assigned a moderate level of confidence. As a quality control step, we also translated all the *lipL32* sequences in frame to confirm that they resulted in functional amino acid sequences. Although the dual indexing and paired end analysis approach described above provides additional confidence in the accuracy of these sequences, we acknowledge that these putative alleles could benefit from further validation. As such, no major phylogenetic conclusions are inferred from these 14 lower confidence alleles and none of these alleles define the novel *lipL32* clades described herein. However, all 34 alleles were used to illustrate the diversity of pathogenic *Leptospira* spp. genotypes present in Puerto Rico soil and water.

### Gene sequencing *secY*

Unlike *lipL32*, which has only been documented in pathogenic leptospires, the *secY* gene is present in both pathogenic (P1 and P2) and saprophytic (S1 and S2) *Leptospira* groups (1, 72). It is also commonly used to obtain *Leptospira* spp. identification from clinical samples. Thus, to compliment the *lipL32* sequence data and provide support for phylogenetic membership to the pathogenic clade and the relationship to other pathogenic leptospires, we generated *secY* sequences for these environmental samples. This was an important quality control step because divergent alleles that have not previously been described from the *lipL32* gene were observed in these samples. Twenty-one *secY*-203bp sequences from 16 samples (3 water and 13 soil) and 29 *secY-*549bp sequences from 23 samples (2 water and 21 soils) were generated (**Table S1**). In combination, *secY* sequences were generated from 32/86 positive samples representing 10/15 sites. All sequences fell within the pathogenic group of the *secY* phylogeny (**Figure S3**), thereby confirming the *lipL32* results; nucleotide sequences are provided in **Table S7**.

### Mixtures

Sequences generated for the *lipL32* and *secY* genes revealed mixtures (multiple pathogenic genotypes) in 39 soil and two water samples from 12 sites; up to five unique *lipL32* genotypes in a single sample were identified (**Table S1 and Table S6**).

### Phylogenetics

Phylogenetic analyses of *lipL32* sequences revealed a high level of genetic diversity among these samples, including four previously undescribed pathogenic *Leptospira* phylogenetic clades (3, 5, 8, and 9; **Figure 2 and Figure S4**); 18 sequences obtained from the environment in Puerto Rico assigned to these four clades. Clades 4 and 6 contain 128 *lipL32* sequences from Puerto Rico as well as environmentally acquired isolates from other locations (1). The two most common *lipL32* genotypes from soil and water in Puerto Rico were a perfect match to *L. gomenensis* in clade 6 (n=56) or a close match to *L. yasudae* (previously *L. dzianensis*) in clade 4b (n=28) (**Figure 2 and Figure S4)**. Interestingly the *L. gomenensis* genotype was recently described in New Caledonia and *L. yasudae (L. dzianensis)* in Mayotte (1), suggesting that these environmental *Leptospira* spp. are distributed globally. The *secY* phylogeny provided additional confidence in the phylogenetic placement of these novel types because those genotypes were also a close match to *L. yasudae* and *L. gomenensis* (**Figure S3**). Interestingly, there also was sequence similarity to *L. santarosai* for some *secY* genotypes, including those from the newly acquired LGVF01 and LGVF02 isolates that are described herein (**Figure S3**). Clades 1, 2, and 7 of the *lipL32* phylogeny (**Figure 2 and Figure S4**) contain described pathogenic leptospires that are commonly found in human and animal infections: *L. interrogans, L. noguchii*, and *L. kirschneri* (clade 1); *L. borgpetersenii* and *L. santarosai* (clade 2); and *L. alexanderi, L. weilii*, and *L. mayottensis* (clade 7). A single sequence from site 5 assigned to clade 1 (**Figure 2 and Figure S4**) and no sequences from Puerto Rico assigned to clades 2 and 7.

**Figure 2:**
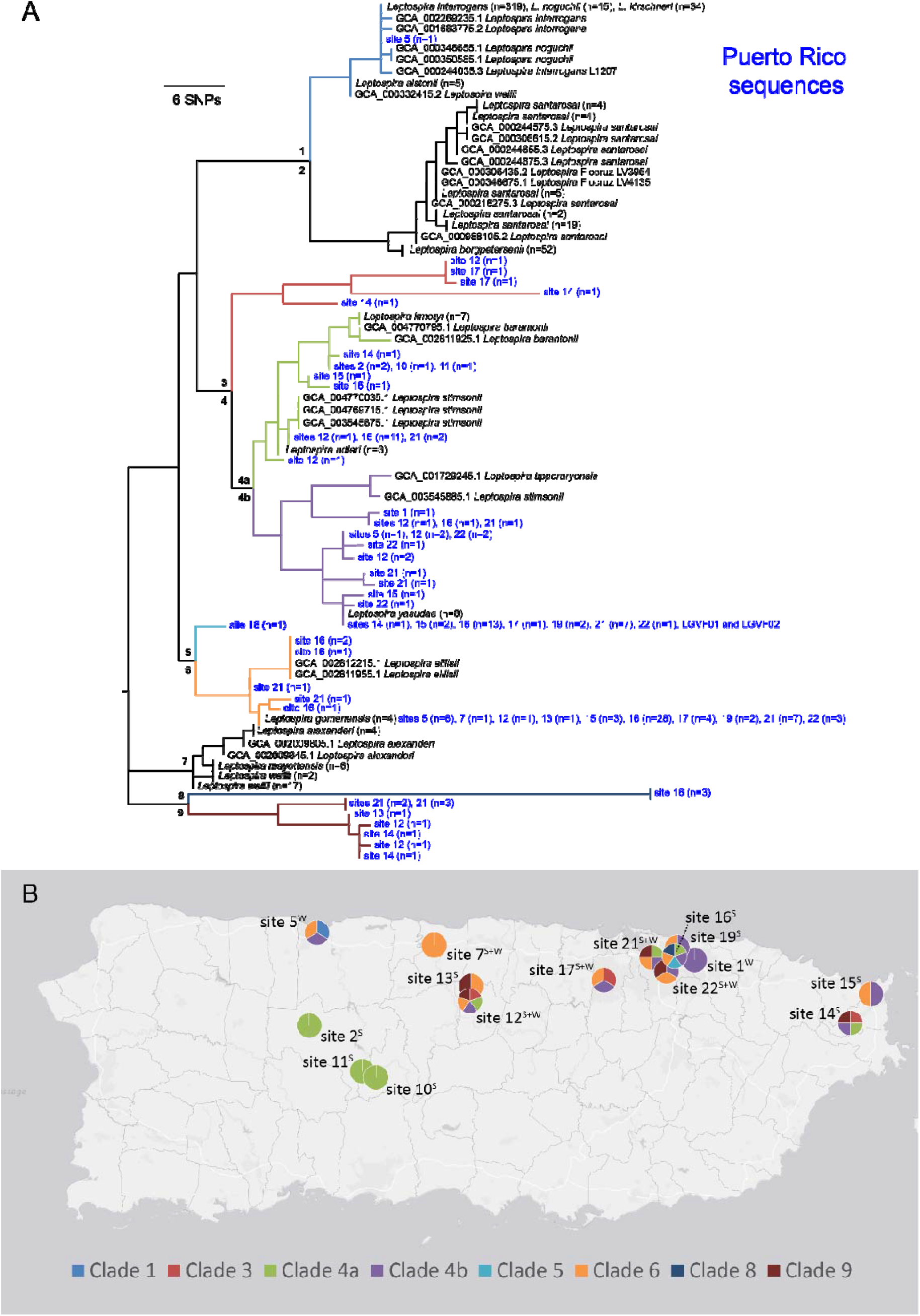
Genetically diverse and previously undescribed lineages of pathogenic *Leptospira* are widespread in Puerto Rico. A) Maximum likelihood phylogeny constructed using a 202bp segment of the *lipL32* gene, including sequences obtained from soil and water samples collected in this study (blue text) and those available in public databases (black text). Nine major genetic clades are represented, seven of which were present in Puerto Rico, including four not previously described. Clades are labelled 1 through 9 on the tree and color coded. B) Sites where each major clade was collected, with colors within the pie charts corresponding to those in the phylogeny; superscript indicates if soil (S) and/or water (W) samples were positive at each site. Fifteen positive sites are included on this map.

### Detection Statistics

We observed a significant association between the distribution (median) of pH values for samples collected from water that were positive (median pH=8.4) and negative (median pH=8.2) for pathogenic *Leptospira* spp. (W=550, *p*=0.0015), but not from soil, where positive and negative median pH values were ~7.5 regardless of pathogenic *Leptospira* status (W=3057, *p*=0.9399). We also detected pathogenic Leptospira spp. in soil more commonly than in water: 48/210 (22.9%) of soil samples compared to 15/210 (7.1%) of water samples (x^2^=15.087, *p*<0.001). No other significant associations were observed among environmental/sampling variables and the detection of pathogenic *Leptospira* spp. (**Table S4**).

### Persistence in soil

During our first sampling period (December 2018) we identified one exploratory site in an urban district of San Juan where seven soil samples (out of ten) tested positive for the presence of pathogenic *Leptospira* spp. DNA (**Table S1**), the highest single site detection rate from this first period. We revisited the site two months later (February 2019) and collected an additional ten soil samples in a linear transect along the edge of the water to test the hypothesis that pathogenic L*eptospira* spp. persists in water-soaked soil at this site; all ten of the new soil samples tested positive for pathogenic *Leptospira* spp. DNA (**Table S1**). We revisited this site once again 14 months after the initial sampling period (February 2020) and collected 20 samples clustered along the edge of the water to use for live *Leptospira* spp. recovery and isolation attempts, as well as persistence analysis; 13 of these 20 soil samples tested positive for pathogenic *Leptospir*a spp. DNA (Table S1). Thus, at all three time points the majority of soil samples collected at this site were positive for pathogenic *Leptospira* spp. DNA: 7/10, 10/10, and 13/20 of samples, respectively (**Figure 3**). Among these positive samples, identical *LipL32* sequences from clades 4b and 6 were identified across all three time points (**Figure 3A)**, suggesting these genotypes persisted in soil at this site.

**Figure 3:**
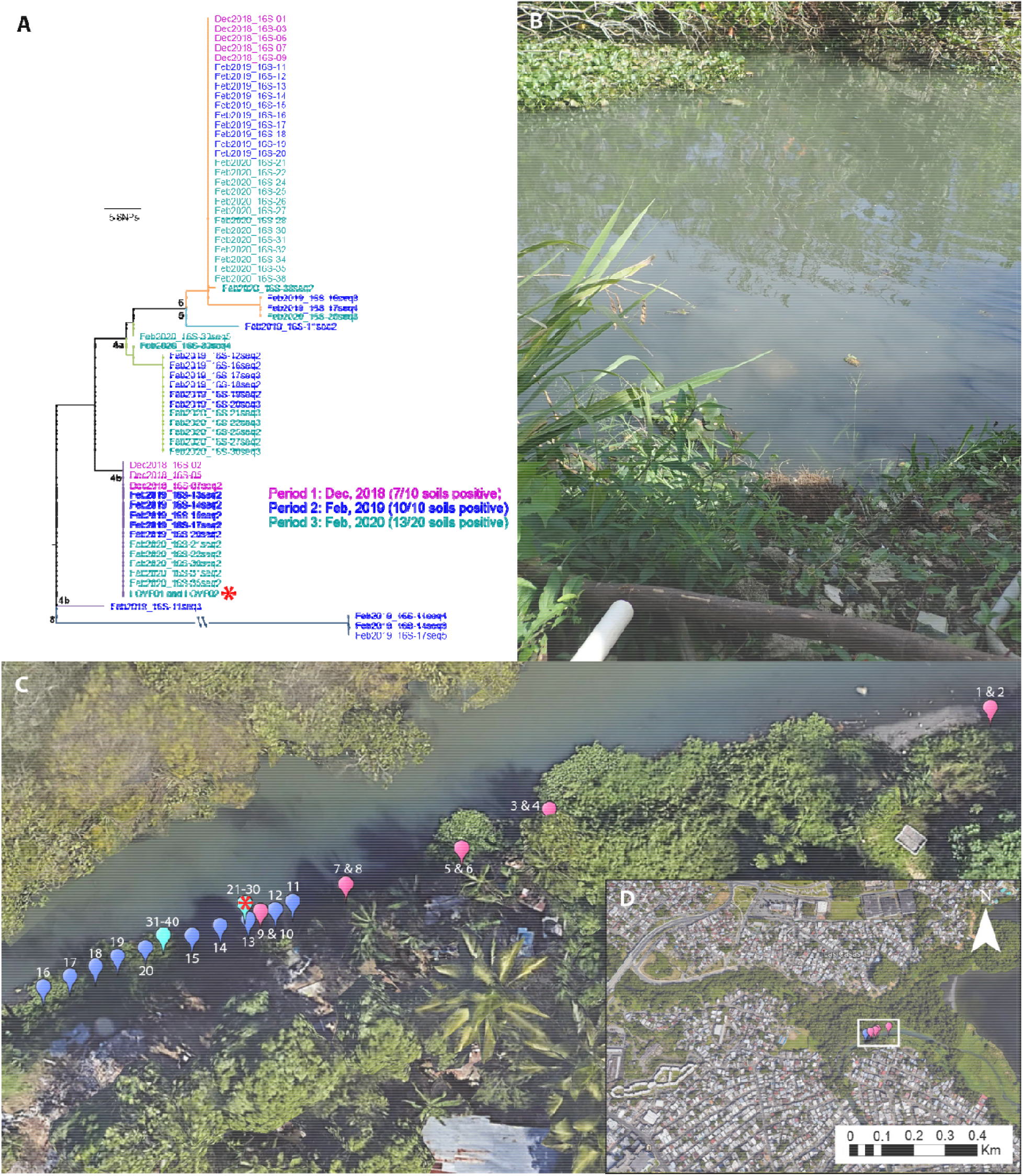
Site 16 in Caño Martín Peña, San Juan, Puerto Rico where sampling was conducted at three separate timepoints to assess environmental persistence of pathogenic *Leptospira* in soil near the edge of the water. A) Maximum likelihood phylogeny of all *lipL32* sequences obtained from this soil site over the course of three sampling periods (color coded by date of sampling period); two of five clades detected at this site were identified at all three timepoints, suggesting environmental persistence. Clade colors correspond to Figure 2. Two isolates representing a novel pathogenic *Leptospira* species were obtained from soil sample 27 (red asterisk). B) Photo of exact location where samples 21-30 were collected and the isolates were obtained. C) Transect where 40 soil samples were collected over three sampling periods. Balloons are color coded to indicate sampling period as in the phylogeny (panel A) and the red asterisk denotes the location where the *Leptospira* isolates originated. D) Location of site 16 in Caño Martín Peña.

### Quantification of pathogenic *Leptospira* spp. in soil cultures

Single aliquots of culture from FAT-positive soil sample #27 and FAT-negative soil sample #40 (both from site 16) were defrosted in a water bath at 37°C and used for a confirmative FAT and for coating onto ELISA plates. As expected, sample #27 presented with a high proportion of LipL32-reactive leptospires (**Figure S5A**), which was quantified by ELISA (**Figure S5B**); approximately 50% of the mixed population was pathogens. In contrast, only basal reactivity was obtained when wells were coated with cells from sample #40, consistent with a small number, or absence of, pathogenic leptospires. Cultures were seeded onto HAN plates and incubated at 37°C in a 3% CO_2_ atmosphere; colonies were typically observed in five days. Colonies were picked from both plates (27 colonies from sample #27 and 54 from sample #40), vigorously homogenized in liquid HAN, and the suspensions used to coat FAT slides. In strong agreement with ELISA results, 14 out of 27 colonies from sample #27 were FAT positive (51.9%), whereas none of the 54 colonies from sample #40 showed significant LipL32 reactivity. Representative FAT images of positive (colony 9 and 17) and negative (colony 8) clonal suspensions from sample #27 are presented in **Figure S5C**. FAT-positive cells were inoculated into liquid HAN and harvested at mid-log phase for PCR confirmation, wherein colony 8 was also employed as a negative control. All 14 FAT-positive cultures were also PCR-positive for both lipL32 and secY and, as expected, no bands were observed when cells from colony 8 were used as a template (**Figure S5D**).

### Sequence identification of pathogenic colonies

All *secY* amplicons were sequenced and the assembled sequences (446bp in length after trimming) were submitted to nucleotide alignment by BLASTN (43). The 14 clones presented an average identity of 95% to *L. santarosai* serovar Princestown strain TRVL 112499. We identified two clones (colony 9 and 18) that were slightly divergent from the other 12 and contained several SNPs within the *secY* sequence (**Figure S3 and Table S7)**. One representative isolate of each genotype, designated as strain LGVF01 and strain LGVF02, respectively, were re-grown in liquid HAN for DNA extraction and whole genome sequencing. As a quality control step we also sequenced the *lipL32* gene from these isolates and confirmed that they matched the sequences we obtained from the originating soil samples.

### Isolation of other leptospires

A total of 40 saprophytic isolates also were obtained during the purification process for pathogenic colonies. We sequenced near full length 16S rRNA fragments (1,330bp) for these isolates using universal primers (73) and identified two genotypes among them that differed by four SNPs. One genotype (16S-01) was a perfect match to L. *mtsangambouensis, L. jelokensis, L. noumeaensis, L. congkakensis, L. levettii*, and *L. macculloughii* and the other (16S-02) was a perfect match to *L. bandrabouensis, L. kemamanensis, L. bouyouniensis, and L. meyeri;* three representatives of each genotype were whole genome sequenced (**Table S8**).

### WGS

In total, we generated whole genome sequences for two pathogenic and six saprophytic isolates obtained from a single soil sample (#27) at the serially sampled site 16 (**Figure 3**); the former are two distinct strains of a novel pathogenic *Leptospira* species that differ from each other by 20,980 pairwise SNPs (4,982 non-synonymous) (**Figure 4B**). This novel pathogen species falls within clade 4b of the *lipL32* phylogeny (**Figure 2** and **Figure S4**) and is most closely related to, but is clearly distinct from, *L. yasudae*; the ANI between these species is 92.8%. Samples from five other positive soil sites and one water site from eastern Puerto Rico (sites 14, 15, 17, 19, 21, and 22) share the same *lipL32* genotype found in this novel pathogenic species, suggesting it may be fairly common and widespread in Puerto Rico (**Figure 2**). The six saprophytic isolates represent three previously known species of Leptospira: they were genomically identified as *L. levettii* (n=1), *L. bandrabouensis* (n=4), and *L. mtsangambouensis* (n=1) (**Figure 4)**. The 16S-01 genotype was conserved among the *L. levettii, L. mtsangambouensis*, and one of the *L. bandrabouensis* isolates, whereas the 16S-02 genotype was conserved among the remaining three *L. bandrabouensis* isolates (**Table S8**). The *L. bandrabouensis* isolate (Sapro03) that contained the 16S-01 genotype differed from the other three *L. bandrabouensis* isolates by >47,000 SNPs (**Figure 4B**). The genome assembly details for all eight isolates are provided in Table S9.

**Figure 4:**
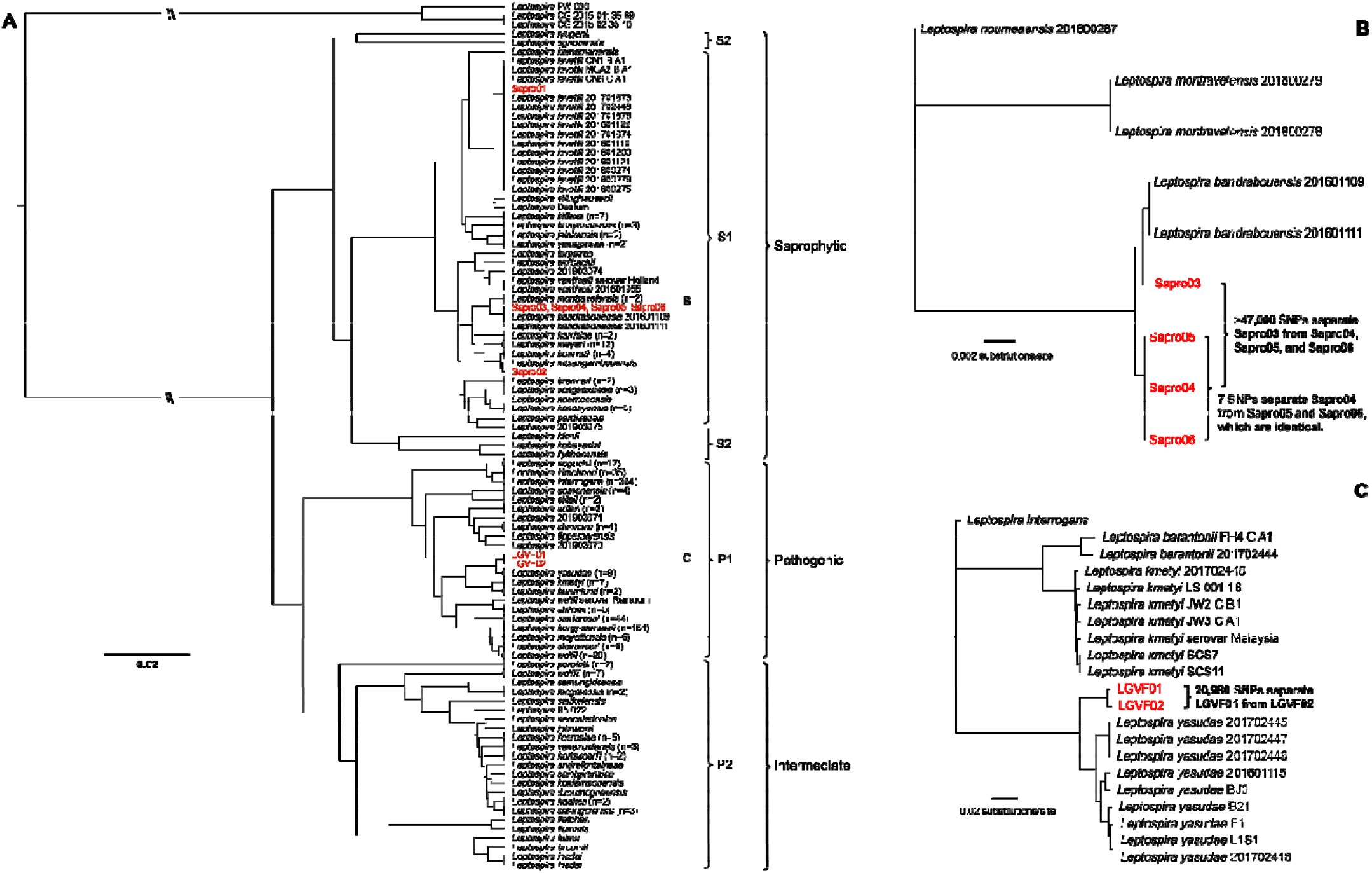
Whole genome dendrogram of 802 known pathogenic, intermediate, and saprophytic *Leptospira* spp. isolates, including six saprophytic and two pathogenic isolates obtained in this study from a single soil sample from site 16. A) Pairwise genomic distance dendrogram contextualizing the relationship among all known saprophytic (S1 and S2), intermediate (P2), and pathogenic (P1) *Leptospira* spp. reveals that the pathogenic isolates LGVF01 and LGVF02 represent two genotypes of the same previously undescribed pathogenic species, whereas the saprophytes belong to three known *Leptospira* spp. B) Detailed view of the S1 clade that contains four saprophytic isolates obtained during this study. This maximum likelihood phylogeny reveals significant diversity among isolates that fall within the *L. bandrabouensis* clade. C) Detailed view of the P1 clade that contains two pathogenic isolates obtained during this study. This maximum likelihood phylogeny reveals two genotypes of a novel pathogenic *Leptospira* spp. isolated from soil in Puerto Rico.

### Hamster infection and MAT response

MAT reactivity was detected when sera from hamsters infected with the new pathogenic isolate LGVF02 were tested against the challenge isolate (LGVF02) at a 1:1600 dilution. However, no experimentally infected hamsters showed clinical signs of infection or weight loss after inoculation (**Table S10**) and at three weeks post-infection all hamster kidneys were culture negative. In addition, at three weeks post-infection all hamsters were seronegative for the 18 serovars, representative of 15 serogroups, listed in **Table S5**.

## Discussion

This study provides new insights into the persistence of pathogenic leptospires in the environment in Puerto Rico and raises important questions regarding host/environmental adaptation, genomic exchange, the evolution or acquisition of pathogenicity in the genus *Leptospira*, and the disease potential of environmental leptospires. Our results also demonstrate the need for updated diagnostics to detect and identify these novel and divergent types.

Our survey for pathogenic *Leptospira* spp. in the environment in Puerto Rico revealed that diverse and novel pathogenic types are widespread in soil and water across the island, and more common in soil than water. Thirty-two previously undescribed *lipL32* alleles were identified, which suggests a vast amount of uncharacterized diversity within the pathogenic group of this genus present in the environment in Puerto Rico. However, we only identified a single sequence from one water sample that shared a *lipL32* genotype with *Leptospira* spp. from clade 1; species from clade 1 are commonly found in human leptospirosis infections. This might suggest that clade 1 types are not adapted for survival in soil and water. Indeed, host adaptation among these commonly identified pathogenic types has been suggested (7), which could explain the limited detection of these pathogenic types in soil and water in this study. This is in line with the described transmission dynamics of human leptospirosis in which infecting bacteria are maintained in the kidneys and renal tubules of animal reservoir hosts and excreted through urine whereby transient contamination of soil and water can occur, leading to human infections. In contrast, we identified 33 less common or novel *lipL32* alleles in these environmental samples whose disease potential is unknown. Although the pathogencity of the novel types is yet to be determined, the presence of the *lipL32* gene suggests pathogenic potential in humans and animals. Of course it is also possible that the opposite is true and that the presence of the *lipL32* gene in these strains is not indicative of pathogenicity but perhaps an artifact of genetic exchange and recombination and/or serves some other purpose in these complex environments. Either way, the abundance and persistence of these types in soil and water suggests adaptation for survival, and potentially proliferation, in these environments.

In support of this hypothesis, we found evidence for *lipL32* positive *Leptospira* spp. persisting in water-soaked soil near the edge of the water for more than one year at an urban site in San Juan, Puerto Rico. Two of the five pathogenic genetic clades found in soil at this site were resampled again at two months and 14 months after the initial sampling date. Although survival and proliferation in this environment seems the most likely explanation, possibly via formation of biofilms (74, 75), it is also possible that this site is continually contaminated from some other source. This same urban site was described in a 2015 study wherein the research team evaluated *Leptospira* seroprevalence in humans and carriage in rodents (76). It was determined in that study that *Leptospira* exposure was high in humans in this urban community (seroprevalence of 27.2%) but the species/serotypes identified in humans and the species identified and/or recovered from rodents in that study were not a match to those found in our study but, rather, were the commonly identified human and animal pathogens *L. borgpetersenii* and *L. interrogans*. However, it is important to point out that seroprevalence testing for this other study employed a MAT panel containing common human and animal pathogenic types only (27 serovars represented), so if exposure from these novel soil dwelling types had occurred they may not have been detected with this method.

Our discovery of so many novel genotypes at this urban site (n=8) and in Puerto Rico as a whole (n=32) highlights the need to address the limited scope of leptospirosis diagnostics (*e*.*g*., a MAT panel that does not include local strains), as it is possible that these novel pathogenic types also are associated with human and animal disease in Puerto Rico but remain undetected in the clinic. Indeed, it is thought that many cases of leptospirosis go undetected or are misdiagnosed as other febrile illnesses due to challenges with implementation and/or limitations of the diagnostic tools themselves (77, 78). If it were shown that these types contribute to human and animal disease and proliferation in the environment is experimentally confirmed, it would contradict the current dogma that pathogenic leptospires require an animal reservoir host for maintenance and proliferation and, thus, suggest a much more complicated scenario of leptospirosis transmission that includes infection by soil dwelling leptospires.

From soil sampled at this urban site in San Juan we isolated a novel pathogenic *Leptospira* spp. (represented by strain LGVF02), which was confirmed to express LipL32 *in vitro*. Sera from infected Golden Syrian hamsters were positive by MAT when using the challenge strain, although no signs of acute disease were observed. Interestingly, sera from challenged hamsters were MAT negative when tested against a panel of 18 commonly tested strains representing 15 serogroups; as such, we suspect that this isolate represents an entirely novel serogroup and, thus, would not be detected using a MAT panel of common isolates. Importantly, the absence of disease symptoms in challenged hamsters does not negate its classification as a pathogen; other pathogenic *Leptospira* spp., including *L. borgpetersenii, L. mayottensis*, and *L. tipperaryensis*, have also failed to cause acute disease in Golden Syrian hamsters (79, 80). LGVF02 was also subjected to LipL32 gene silencing via CRISPRi as part of another study (56) and the results of that study (specifically, the detection of the LipL32 and LipL41 proteins) further support the pathogenic classification of this novel leptospire. The species description of LGVF02 will be treated in a separate publication in which we will investigate whether it represents an entirely new serogroup. Either way, the ability of strain LGVF02 to evade detection with MAT based on common isolates highlights the need to include regionally relevant isolates when conducting MAT in the clinic to ensure detection of these lesser known types and better understand their potential role in human and animal disease.

Our attempts to sequence two separate secY amplicons (*secY-*203bp and *secY*-549bp) from *lipL32* positive samples suggested that SNPs or other genetic mutations were present in the PCR priming sites. We suspect this because *secY* PCR amplification was only achieved in 32 of 86 (37%) *lipL32* positive environmental samples from Puerto Rico. Further evidence of this possibility is supported by other investigators; during the preliminary genome analysis of a recently acquired *L. kirschneri* isolate it was discovered that the reverse priming site for the *secY*-549bp PCR was missing entirely (Camila Hamond, NVSL, personal communication). Accordingly, this *L. kirschneri* isolate and originating sample failed to amplify with *secY*-549bp PCR. The poor performance of both *secY* PCRs with these environmental samples that displayed diverse *lipL32* genotypes, paired with the MAT evasion of the LGVF02 isolate described herein, again highlights the need for updated diagnostics that can account for this novel diversity. However, this goal requires the acquisition of genomes to first characterize this diversity in order that improved diagnostic targets can be developed.

We generated whole genome sequences for two pathogenic and six saprophytic *Leptospira* isolates obtained from a single soil sample collected from serially sampled site 16. The whole genome dendrogram contextualized the phylogenetic relationships of these isolates to all other known *Leptospira* spp. and revealed that the two pathogenic isolates, LGVF01 and LGVF02, were unique strains (separated by 20,980 SNPs) representing a single, novel pathogenic species. We classified these isolates as pathogenic based upon the presence of the *lipL32* gene in their genomes and the expression of the LipL32 protein, as well as their phylogenetic relationships to other species. The saprophytic isolates represent three known species, of which two (*L. bandrabouensis* and *L. mtsangambouensis*) have previously only been identified from the island of Mayotte in the Indian ocean, and the third (*L. levettii*) has previously been identified only in Mayotte, Malaysia, and New Caledonia. In other words, these three species have not previously been identified in Puerto Rico, the Caribbean, or the Western Hemisphere. In total, four *Leptospira* species were isolated from a single 10g soil sample from site 16 in Puerto Rico, and the *lipL32* analysis from this same site identified four additional phylogenetic clades that were not represented by the isolates obtained from this sample (**Figure 3A**), suggesting the presence of even more pathogenic species in soil from this site. Finally, the SNP diversity identified among isolates within the same species from a single soil sample is considerable (20,980 between LGVF01 and LGVF02 and >47,000 SNPs differentiating two subgroups of *L. bandrabouensis*). Together, these findings reveal a tremendous amount of unrecognized *Leptospira* spp. diversity within the environment in Puerto Rico, and very likely many other geographic locations, that has yet to be characterized.

Soil may serve as a long-term reservoir for contamination of waterways with pathogenic *Leptospira* spp. in Puerto Rico. At four of the five sites where pathogenic leptospires were detected in both soil and water, the *Leptospira* clade(s) identified in water were a subset of those identified in soil and a higher proportion of soil samples were positive when compared to water samples at these sites. These observations suggest soil may serve as a reservoir for pathogenic *Leptospira* that leads to the contamination of waterways in Puerto Rico, which could occur when the soil is disturbed by animal or human activity, or during weather events such as heavy rains and floods. This is in line with the dynamics of *Leptospira* environmental contamination reviewed elsewhere (28). However, our findings also provide support for the hypothesis that some pathogenic *Leptospira* spp. are maintained in the soil long term, rather than just transiently being present due to urine contamination from an infected reservoir host. If this is the case, the potential for genetic exchange among pathogenic isolates that could lead to the emergence of novel outbreak strains is something that needs to be explored further. A possible example of this may be the documented outbreak between 1999-2003 in Thailand that was associated with a novel type of *L. interrogans* that emerged, caused illness in humans for several years, and then inexplicably seemed to fade out (81). It is possible this type of outbreak is due to the exchange of genetic material among transiently present (via urine contamination) and soil dwelling pathogenic leptospires that result in increased pathogenic potential of certain strains in these complex environments. A comprehensive comparative genomics study conducted by Xu *et al*., 2016 (7) revealed that horizontal gene transfer events resulting in the acquisition of novel genes in *Leptospira* are fairly common.

Overall, this study provides evidence for the existence of divergent lineages of soil dwelling pathogenic *Leptospira* spp. in Puerto Rico; the disease potential of these novel types in humans and animals is unknown. The genetic diversity within this genus found in the environment in Puerto Rico is vast and, thus, there is a need to characterize this diversity to account for it in the diagnosis of leptospirosis. Many cases of leptospirosis are thought to go undetected, or are misdiagnosed as other febrile illnesses, and it is possible that these novel types play a role in such cases. A follow up study in Puerto Rico to investigate the possibility that these novel types are found in human and animal samples would be highly valuable. Furthermore, environmental surveys aimed at the discovery of these novel types in all areas where leptospirosis is a public health concern will be of critical importance towards improving diagnostics and public health outcomes.

## Data Availability

All data produced in the present study are available upon reasonable request to the authors

## Acknowledgements

We thank Ryelan McDonough and Amber Jones for laboratory assistance preparing samples for whole genome sequencing and Karen LeCount for performing MAT. USDA is an equal opportunity provider and employer. Mention of trade names or commercial products in this publication is solely for the purpose of providing specific information and does not imply recommendation or endorsement by the U.S. Department of Agriculture or the Centers for Disease Control and Prevention. This work was funded in part by the United States Department of Health and Human Services Centers for Disease Control and Prevention (Award# 6NU1ROT000006-01-04). The Brazilian agency FAPESP also financially supported this work; LGVF is funded with a fellowship from FAPESP (2017/06731-8 and 2019/20302-8).

## Supporting information

**Figure S1:**
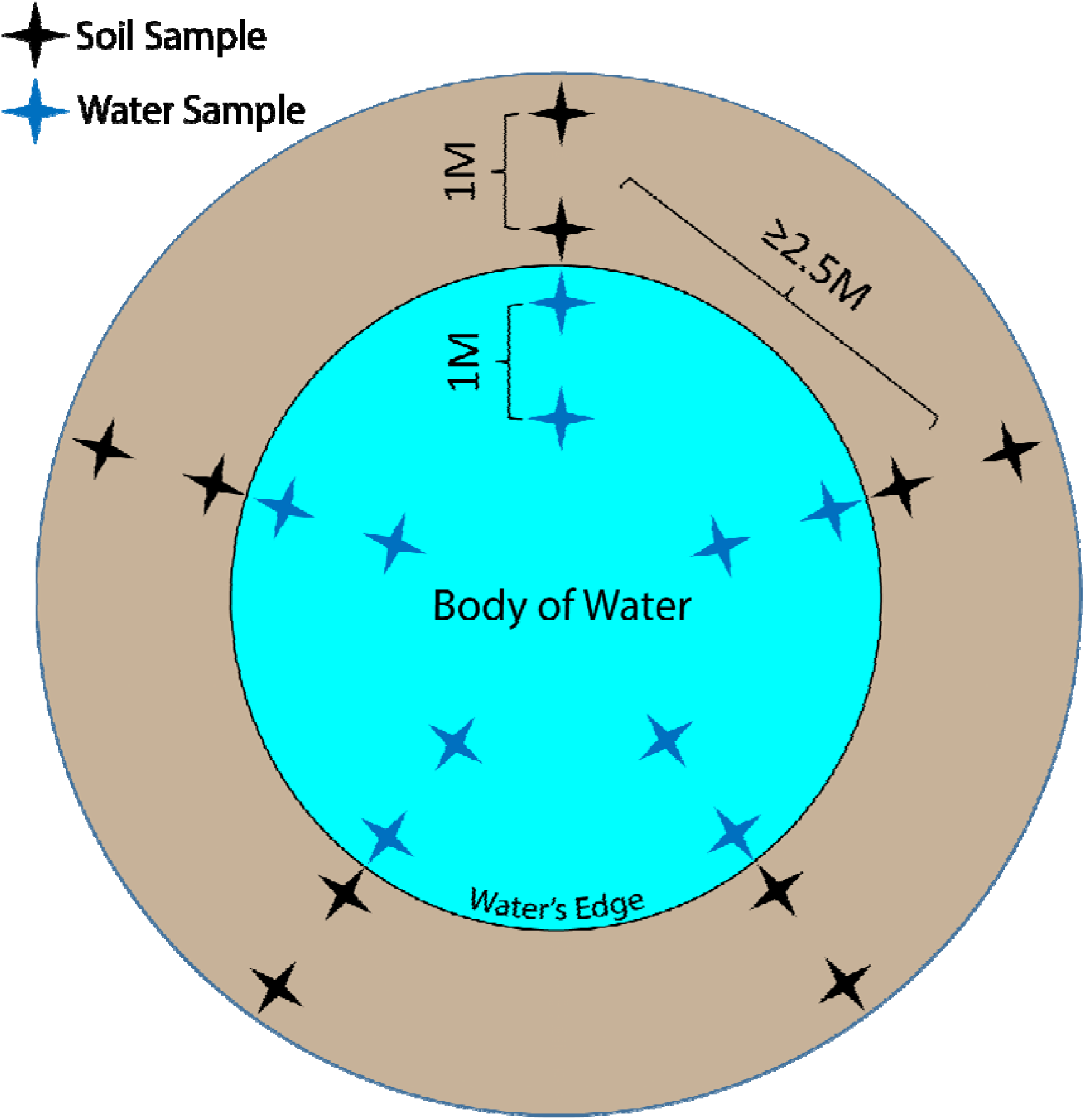
Sampling design for paired soil and water sites. At each site, five transects were sampled (two soil and two water samples per transect) for a total of 20 samples per site. Black and blue stars represent soil and water samples, respectively. Transects were a minimum of 2.5 meters apart. This sampling design was also applied in a linear fashion for the sampling of rivers and streams.

**Figure S2:**
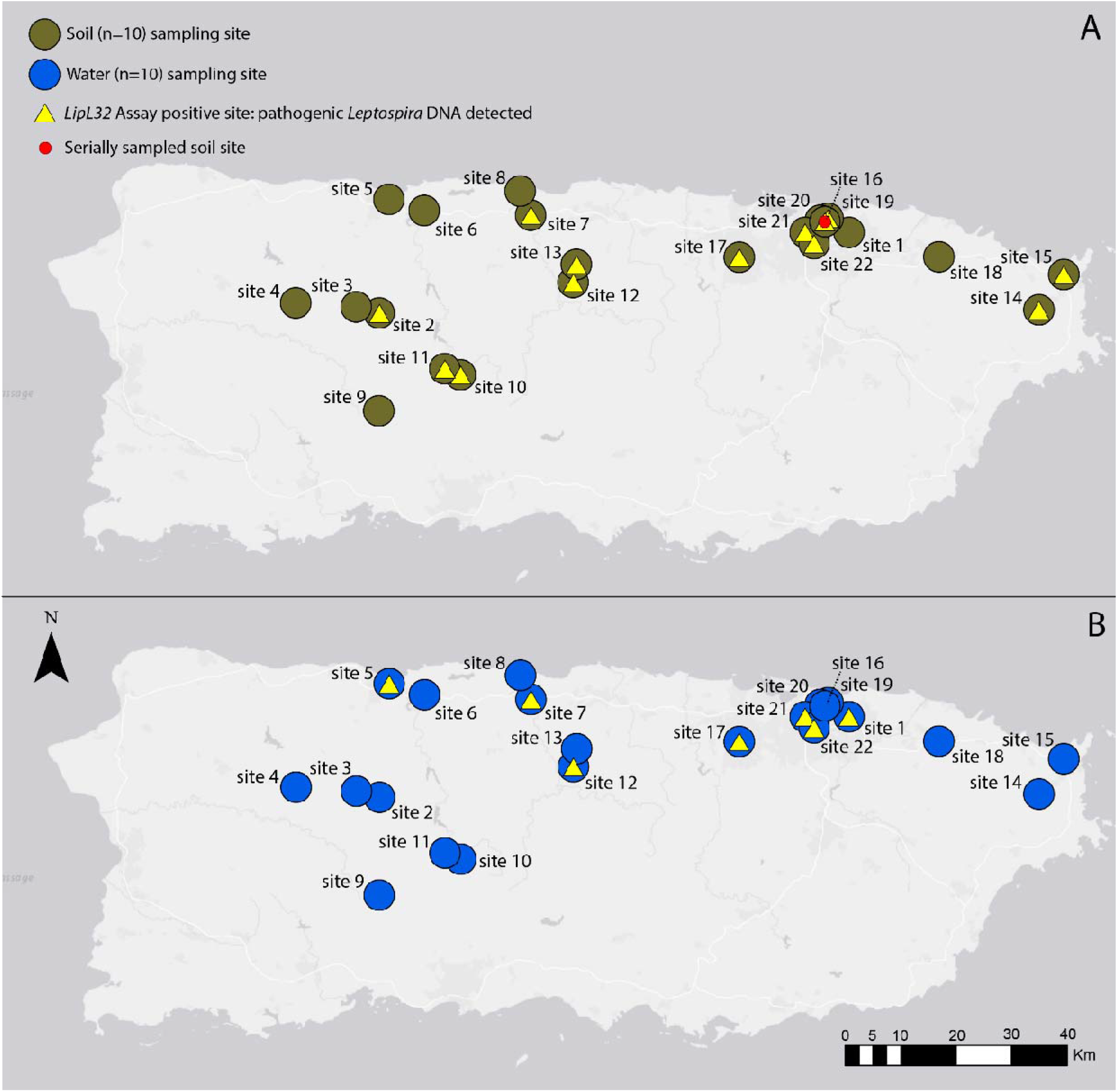
Map of Puerto Rico indicating where soil (panel A) and water (panel B) samples were collected for the detection of pathogenic *Leptospira* spp. (2018-2020). The yellow triangles indicate sites where pathogenic *Leptospira* DNA was detected. Eight soil sites were positive for pathogenic *Leptospira* DNA, compared to two water sites; both soil and water were positive at five sites. Site 16, which was used to assess environmental persistence in soil is indicated with a red circle.

**Figure S3:**
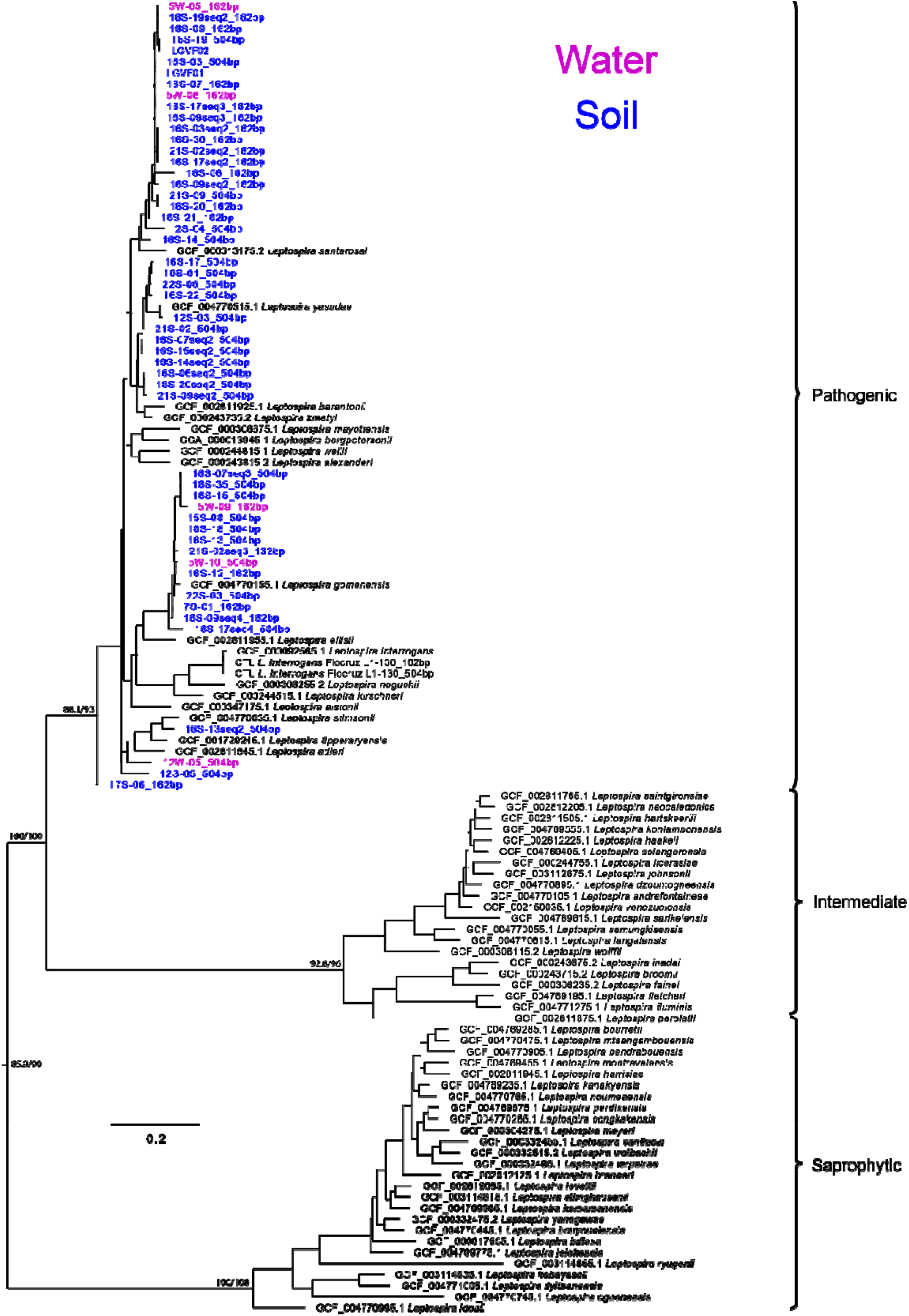
Maximum likelihood phylogeny of the *secY* gene reveals that pathogenic *Leptospira* was identified in Puerto Rico soil and water and provides support for the phylogenetic relationships observed with *lipL32* sequences. Sequences generated from soil and water samples collected in Puerto Rico are in blue or pink text, the novel pathogenic Puerto Rico isolates from soil are also included; reference sequences for pathogenic, intermediate, and saprophytic *Leptospira* are in black. Bootstrap/aLRT support values are indicated on branch nodes.

**Figure S4:**
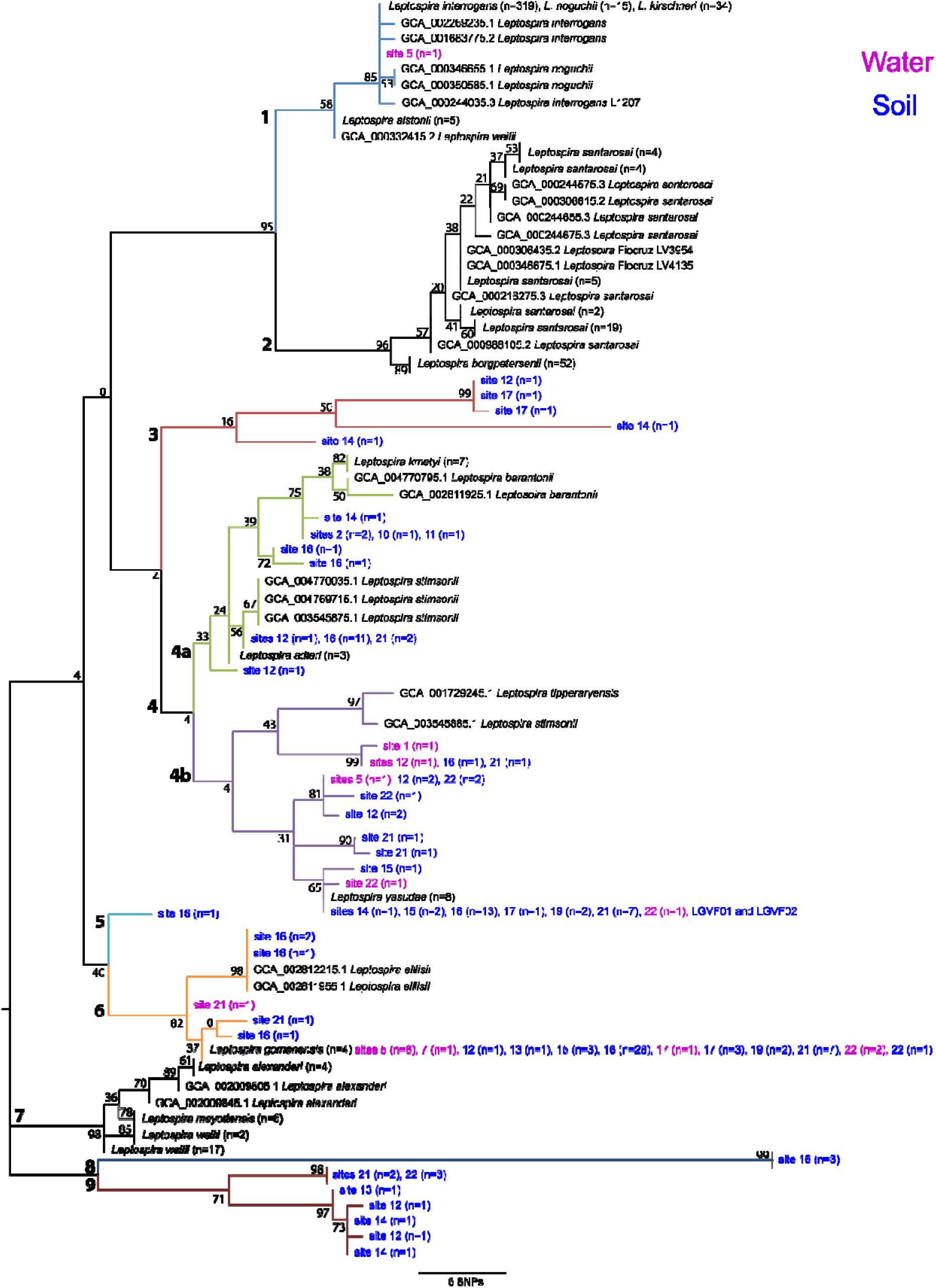
Maximum likelihood phylogeny built using a 202bp region of the *lipL32* gene. Sequences generated from soil (blue text) and water (pink text) samples collected in Puerto Rico are displayed. Reference sequences representing all other known pathogenic *Leptospira* spp. are in black. Nine major clades are represented of which seven were identified in Puerto Rico and four of those (3, 5, 8, and 9) have not been described previously. Identical genotypes were present in soil and water from Puerto Rico at two sites (17 and 21), with both occurrences representing a single genotype from clade 6 (in orange). Bootstrap values are indicated on each branch. Major clades are numbered 1 through 9 and color-coding matches Figure 2.

**Figure S5:**
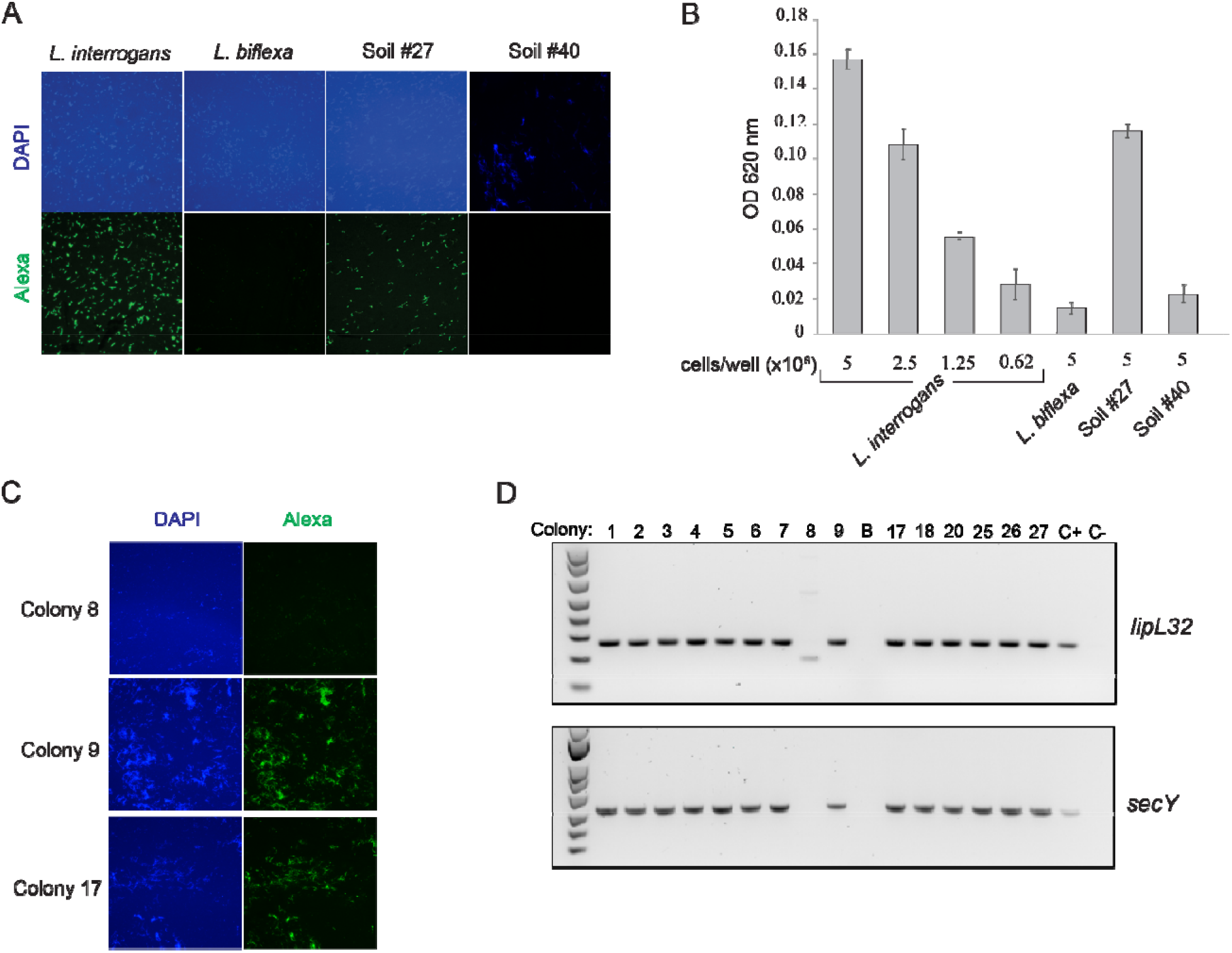
Identification and isolation of pathogenic *Leptospira* spp. from Puerto Rico soil at serially sampled site 16. *LipL32* PCR positive (Soil #27) and negative (Soil #40) samples were subjected to FAT (Panel A) and ELISA (Panel B) along with positive and negative controls (*L. interrogans* and *L. biflexa*, respectively) to assess expression of the LipL32 pathogenicity protein. Individual colonies from Soil #27 were subjected to additional FAT testing (Panel C) and confirmatory PCR (Panel D) to verify that the obtained isolates were pathogenic.

**Supplemental File 1: Table S1 Results summary**. Summary of detection and sequencing results for environmental samples from Puerto Rico, 2018-2020.

**Supplemental File 1: Table S2 Sampling data**. Sample and site ID, location, elevation, and pH.

**Supplemental File 1: Table S3 Accession numbers**. GenBank accession numbers for all sequences used to build the *lipL32, secY*, and whole genome phylogenies.

**Supplemental File 1: Table S4 Statistical tests**. Statistical tests used to assess associations between the detection of pathogenic *Leptospira* spp. and environmental or sampling variables.

**Supplemental File 1: Table S5 MAT panel**. Antigens used in the microscopic agglutination test (MAT) that were unreactive when tested against sera from hamsters infected with the novel pathogenic *Leptospira* spp. isolated from Puerto Rico soil.

**Supplemental File 1: Table S6 *lipL32* sequences**. All *lipL32* sequences generated during this study using Sanger and AmpSeq methods.

**Supplemental File 1: Table S7: *secY* sequences**. All *secY* sequences generated during this study for two *secY* amplicons (203 and 549bp).

**Supplemental File 1: Table S8: 16S sequences**. Near full length 16S sequences (1330bp) generated for six saprophytic isolates.

**Supplemental File 1: Table S9: Genome assembies**. Genome assembly details including assembly size, number of contigs, and NCBI assembly accession numbers.

**Supplemental File 1: Table S10: Hamster infection**. The weight of each hamster was recorded daily, beginning two days preinfection.

